# Clinical and epidemiological characteristics of Madariaga and Venezuelan equine encephalitis virus infections

**DOI:** 10.1101/2024.02.02.24302220

**Authors:** Luis Felipe Rivera, Carlos Lezcano-Coba, Josefrancisco Galué, Xacdiel Rodriguez, Yelissa Juarez, William M. de Souza, Zeuz Capitan-Barrios, Anayansi Valderrama, Leyda Abrego, Hector Cedeño, Carmela Jackman, Jesse J. Waggoner, Patricia V. Aguilar, Hilda Guzman, Scott C. Weaver, Robert B. Tesh, Sandra López-Vèrges, Christl A. Donnelly, Cassia F. Estofolete, Mauricio L. Nogueira, Nuno R. Faria, Nikos Vasilakis, Amy Y. Vittor, Darci R. Smith, Jean-Paul Carrera

**Affiliations:** Instituto Conmemorativo Gorgas de Estudios de la Salud, Panama City, Panama; Carson Centre for Research in Environment and Emerging Infectious Diseases, La Peñita, Darién, Panama; University of Kentucky, College of Medicine, Lexington, Kentucky, USA; Universidad de Panamá, Ciudad de Panamá; Ministry of Health, Panama City, Panama; Emory University, Atlanta, Georgia, USA; The University of Texas Medical Branch, Galveston, TX, USA; University of Oxford, Oxford, United Kingdom; Faculdade de Medicina de São José do Rio Preto, São José do Rio Preto, São Paulo Brazil; Imperial College London, London, United Kingdom; Faculdade de Medicina da Universidade de São Paulo, São Paulo, Brazil; University of Florida, Gainesville, Florida, USA; Naval Medical Research Command, Ft. Detrick, MD, USA

**Keywords:** Madariaga Virus, Venezuelan equine encephalitis, alphaviruses, outbreak investigation

## Abstract

Madariaga virus (MADV) and Venezuelan equine encephalitis virus (VEEV) are emerging arboviruses affecting rural and remote areas of Latin America. However, there are limited clinical and epidemiological reports available, and outbreaks are occurring at an increasing frequency. We addressed this gap by analyzing all the available clinical and epidemiological data of MADV and VEEV infections recorded since 1961 in Panama. A total of 168 of human alphavirus encephalitis cases were detected in Panama from 1961 to 2023. Here we describe the clinical signs and symptoms and epidemiological characteristics of these cases, and also explored signs and symptoms as potential predictors of encephalitic alphavirus infection when compared to those of other arbovirus infections occurring in the region. Our results highlight the challenges clinical diagnosis of alphavirus disease in endemic regions with overlapping circulation of multiple arboviruses.

## Introduction

Arthropod-borne viruses (arboviruses) infect humans worldwide and cause significant morbidity and mortality. During the past few decades, the emergence and/or resurgence of arboviruses has been increasing to near pandemic proportions and poses a significant global health threat (1). United States (U.S.) military personnel are frequently stationed in areas where these viruses are endemic or where they may emerge, which could threaten military readiness. Latin America is known to be endemic for several arboviruses that pose significant public health concerns. Important alphaviruses in neotropical regions that can cause human disease include Mayaro virus (MAYV), Venezuelan equine encephalitis virus (VEEV), eastern equine encephalitis virus (EEEV), Madariaga virus (MADV; previously considered a subtype of EEEV) and chikungunya virus (CHIKV), while important flaviviruses include dengue virus (DENV), yellow fever virus (YFV) Zika virus (ZIKV), and St. Louis encephalitis virus (SLEV) (2, 3).

VEEV is widely distributed throughout the Americas and at least 14 subtypes and varieties have been described (4). VEEV subtypes IAB and IC can cause explosive, large-scale epizootics in horses and spillover epidemics in humans (5, 6). VEEV enzootic subtypes (i.e., VEEV ID, IE) are associated with a regular incidence of human infections by spillover from enzootic cycles that involve rodents and sylvatic mosquitoes. Evidence suggests that equine-adaptive or mosquito-adaptive mutations in the VEEV enzootic subtype ID led to the emergence of epizootic/epidemic VEEV subtypes (5). VEEV enzootic/endemic subtype ID infection is highly prevalent in the eastern province of Darien, Panama, where human infections are sometimes fatal and seroprevalence in some villages is up to 75% of the population (5). Eastern equine encephalitis virus (EEEV) was reclassified as two different species in 2010: EEEV in North America and MADV in Latin America (7). MADV was not associated with human outbreaks before 2010 when the first known human outbreak was reported in Darien, Panama (8). Both MADV and VEEV circulated simultaneously during this outbreak with 99 acute cases and 19 hospitalizations for encephalitis. Confirmed cases included 13 for MADV, 11 for VEEV, and one case of co-infection. A fatal MADV infection was confirmed in this same region in 2017. Modelling of 2012 and 2017 Darien Province serosurvey data suggested that alphavirus transmission is endemic in this region (9). Many alphavirus disease cases appear to present as a self-limited febrile illness, but persistent neurological signs and symptoms have been reported for up to five years following MADV and VEEV exposure (5).

VEEV and MADV infections are likely underdiagnosed due to the lack of available diagnostic tools and the inability to clinically differentiate them from other arboviral diseases. It is estimated that up to 10% of dengue cases in Central and South America may actually be due to VEEV (10). Complicating this further is the increasing trend in dengue incidence over the last several decades (11). CHIKV and ZIKV had not previously circulated within the Western Hemisphere until they emerged explosively in 2013 and 2014, respectively. Both viruses became endemic in Latin America where they now co-circulate in DENV-endemic regions (3). The clinical presentation of these arboviral diseases can range from asymptomatic or undifferentiated mild febrile illness to severe disease (3).

The increasing geographical spread and disease incidence of arbovirus infections in the Americas is a major public health concern. Undifferentiated febrile illnesses remain a diagnostic and therapeutic challenge in arbovirus-prone regions, due to the lack of available tools for the high diversity of pathogens responsible for these clinical syndromes. MADV and VEEV have been associated with severe or even fatal outcomes, but early after disease onset, these infections are often clinically indistinguishable from other arboviral syndromes, delaying prompt care for patients at risk for more serious outcomes. Here we describe the clinical signs/symptoms and epidemiological characteristics of all reported MADV and VEEV human infections occurring in Panama from 1961-2023. Additionally, we explore potential symptoms as predictors of encephalitic alphavirus infection when compared to those occurring from other arbovirus infections endemic to the region.

## Materials and Methods

### Ethics statement

The use of human data and samples from outbreaks was approved by the Panamanian Ministry of Health (protocol number 2077, protocol: 365/CBI/ICGES/2023, approved on November 30, 2023), and the Gorgas Memorial Institute (GMI) IRB (protocol: 335/CBI/ICGES/21, protocol: 073/CBI/ICGES/21 and protocol 138/CBI/ICGES/22, approved on March 19, 2021).

### Alphavirus surveillance

Upon suspecting MADV or VEEV (henceforth called encephalitic alphavirus infection), health center clinicians submit blood samples to Instituto Conmemorativo de Gorgas de Estudios de la Salud (ICGES), which serves as the National Reference Center for Infectious Disease Diagnostics in Panama. Alphavirus infections are also often identified through the National Dengue Surveillance system or during encephalitis outbreak response activities. The former system, instituted in 1988, initially provided centralized testing of dengue suspected cases submitted by clinicians (1993 – 2009), but subsequently established diagnostic capacity in all local clinical units (12). Some alphavirus infections were identified upon ruling out DENV cases. In addition, several alphavirus outbreak investigations have been conducted since 2010 and consist of community-wide febrile surveillance and serosurveys.

### Alphavirus outbreak case definition

The case definition of a suspected alphavirus encephalitis case included fever and headache, while a probable case was defined as a suspected case with neurological manifestations (e.g., somnolence, lethargy, or seizures). A confirmed case was defined as a suspected or probable case with laboratory confirmation (i.e., viral isolation, RT-PCR, IgM ELISA, or IgG ELISA or PRNT seroconversion of paired clinical samples). The general laboratory algorithm for diagnosis is depicted in Figure 1 and detailed information of laboratory procedures is provided in the supplemental material.

**Figure 1.**
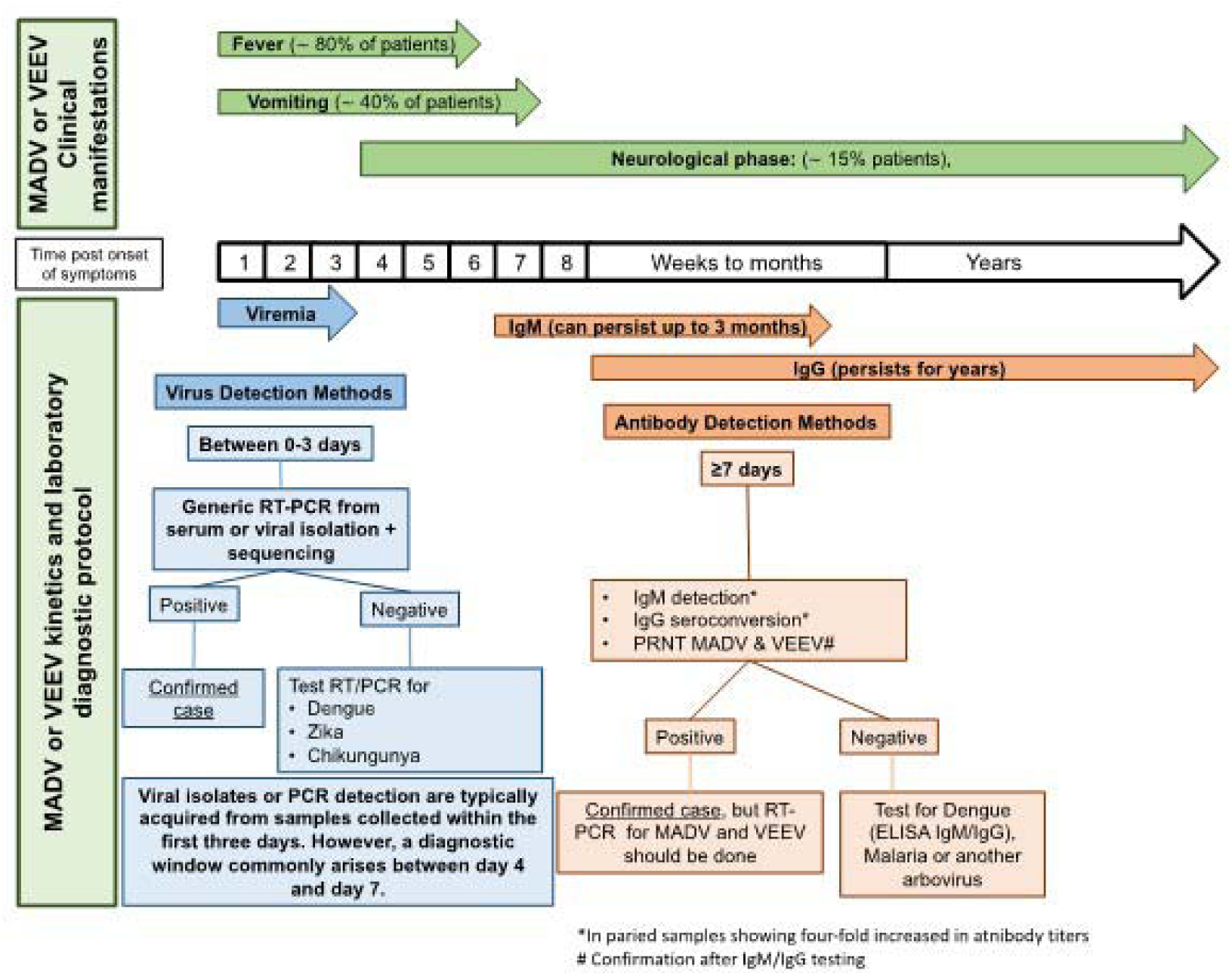
Laboratory algorithm for Madariaga and Venezuelan equine encephalitis diagnosis. Diagnostic algorithm used for two endemic encephalitic alphaviruses based on days since symptom onset.

### Alphavirus data collection

We retrospectively searched and retrieved clinical and epidemiological information of all MADV and VEEV infections reported in clinical records and epidemiological forms from 1961 to 2020, available at ICGES, and extending data published previously (13). Cases detected from 2021 to 2023, were collected as part of the surveillance initiative undertaken by the USA-National Institute of Allergy and Infectious Diseases, Centers for Research in Emerging Infectious Diseases Network initiative. The Coordinating Research on Emerging Arboviral Threats Encompassing the Neotropics (CREATE-NEO) in Panama and the Armed Forces Health Surveillance Division (AFHSD), Global Emerging Infections Surveillance (GEIS) Branch, ProMIS ID P0052_23_NM undertakes acute febrile surveillance across the country. The dataset includes demographic characteristics, clinical symptoms, severity of infection, and sick contacts. When available, geographic coordinates of alphavirus-positive households were also collected. Duplicate or similar signs and symptoms were condensed into composite variables, which provided a better representation of the symptomatology. These were then used to compare clinical manifestations across the main arboviral infections in Panama (MADV, VEEV, DENV, CHIKV, ZIKV).

### Comparison of arboviral symptoms

To account for low statistical power, confirmed MADV and VEEV infections were grouped into a single category. Encephalitic alphavirus cases were defined as all laboratory-confirmed alphavirus infections reported in Panama from 1961 to 2023. Encephalitic alphavirus infections were compared to DENV, ZIKV and CHIKV. A DENV dataset was obtained from a cross-sectional study in 2009 and a ZIKV dataset in 2016. Both DENV and ZIKV datasets were provided by the São José do Rio Preto Health Service in São Paulo State, Brazil, and were published elsewhere (14). The CHIKV data were obtained from CHIKV surveillance in the state of Amazonas, and the City of Recife, Pernambuco, Brazil, from 2015-2020 (15). Additional details of source, laboratory procedures and information of controls are provided in the supplemental material. A flow chart of the alphavirus and endemic arbovirus infections used in this study is depicted in Supplemental Figure 1.

### Statistical methods

Initially, a total of 121 variables associated with participants’ symptomatology were included in the database. These variables underwent categorization and grouping based on specific clinical criteria for each virus. Data were reduced using exploratory factor analysis (EFA) and Principal component analysis (PCA). Variables with zero variance were excluded, using a Kaiser-Meyer-Olkin threshold of 0.6 (Supplemental Table 1). Ultimately, sign and symptoms were reduced to 14 variables, which were used in the analysis.

To evaluate MADV- and VEEV-associated signs and symptoms, we conducted multivariable logistic regression analysis, controlling for age and biological sex. Univariate logistic regressions were undertaken to evaluate the symptoms associated with alphavirus infection (MADV and VEEV) and those reported in DENV-, ZIKV- and CHIKV infections. Variables were selected using a nested log-likelihood ratio test. Variables with more than 10% missing data were excluded from the final analysis. The associations between specific symptoms and viral infection were expressed as odd ratios. A p-value of <0.05 was considered statistically significant. Statistical analyses were undertaken using Stata v.17 and R Studio statistical packages (Statacorp, College Station, TX).

## Results

### MADV and VEEV epidemiology

Between 1961 and 2023, Panama recorded 168 laboratory confirmed MADV and/or VEEV infections. For VEEV infections, there were 131 confirmed cases, with 60 (45.8%) detected during outbreaks and 71 (54.2%) identified through arbovirus surveillance (Figure 2A). For MADV infections, 37 were confirmed cases, with 34 (91.9%) identified during outbreaks and 3 (8.1%) detected through passive arbovirus surveillance (Figure 2B). The socio-demographic characteristics of these cases are described in Table 1. Detailed clinical and epidemiological information was accessible for 132 out of 168 (78.6%) human alphavirus encephalitis infections, comprising 36 MADV infections (27.2%) and 96 VEEV infections (72.7%). The breakdown of age distribution for MADV cases revealed that 16 (44.4%) were in the 0–5-year-old age group, 11 (30.6%) in the 6–20-year-old age group, and 9 (25.0%) were in the ≥21-year-old age group. In VEEV infections, 16 (18.0%) cases were in the 0-5-year-old age group, 35 (39.3%) were in the 6–20-year-old age group, and 38 (42.7%) were in the ≥21-years old age group. Seven cases did not record age information (Table 1).

**Figure 2.**
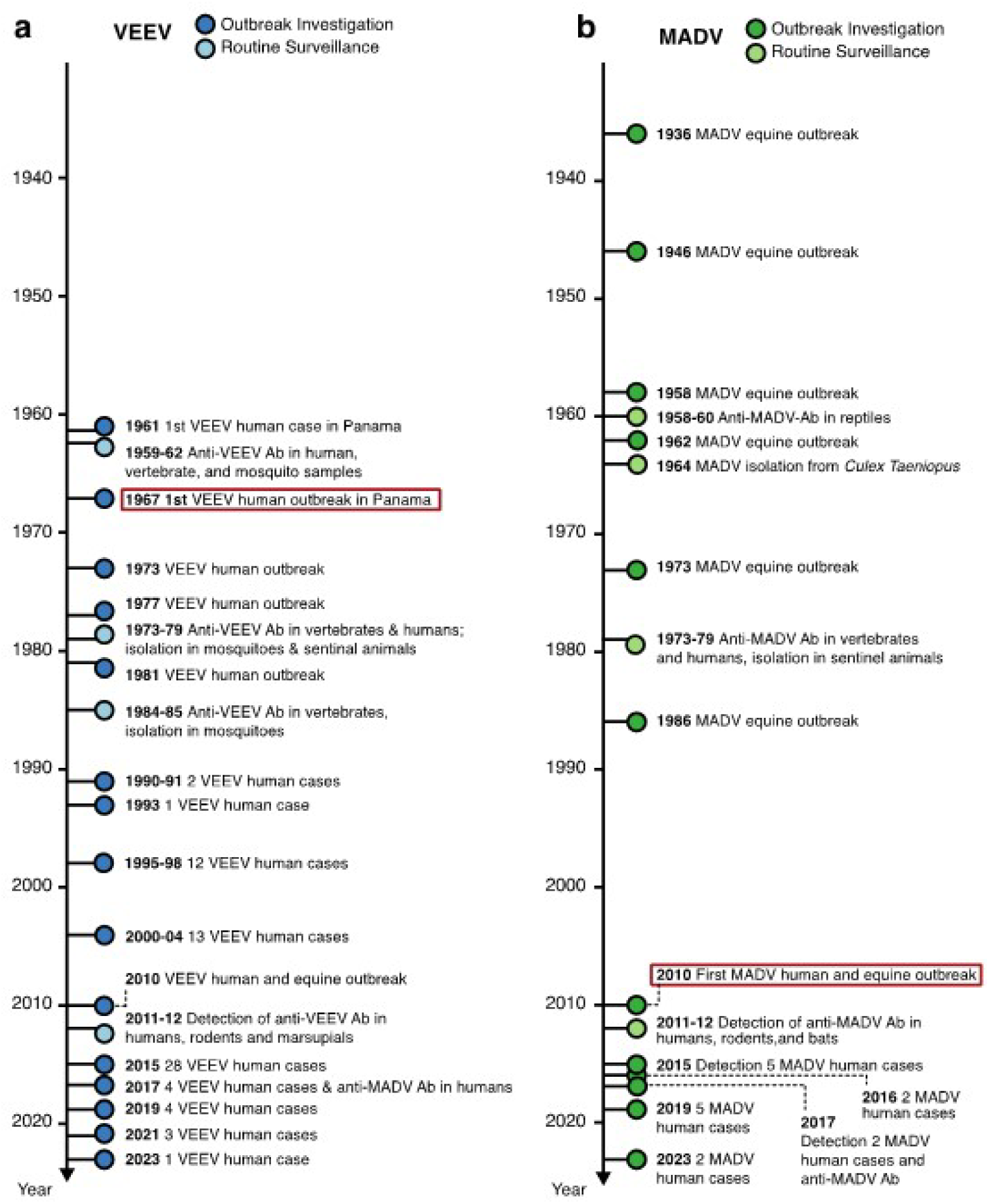
Timeline of recorded a) VEEV and b) MADV important events in Panama. Timeline showing historical of VEEV and MADV events in, human cases, vectors, and animals in Panama over time from 1961 to 2023.

**Table 1.**
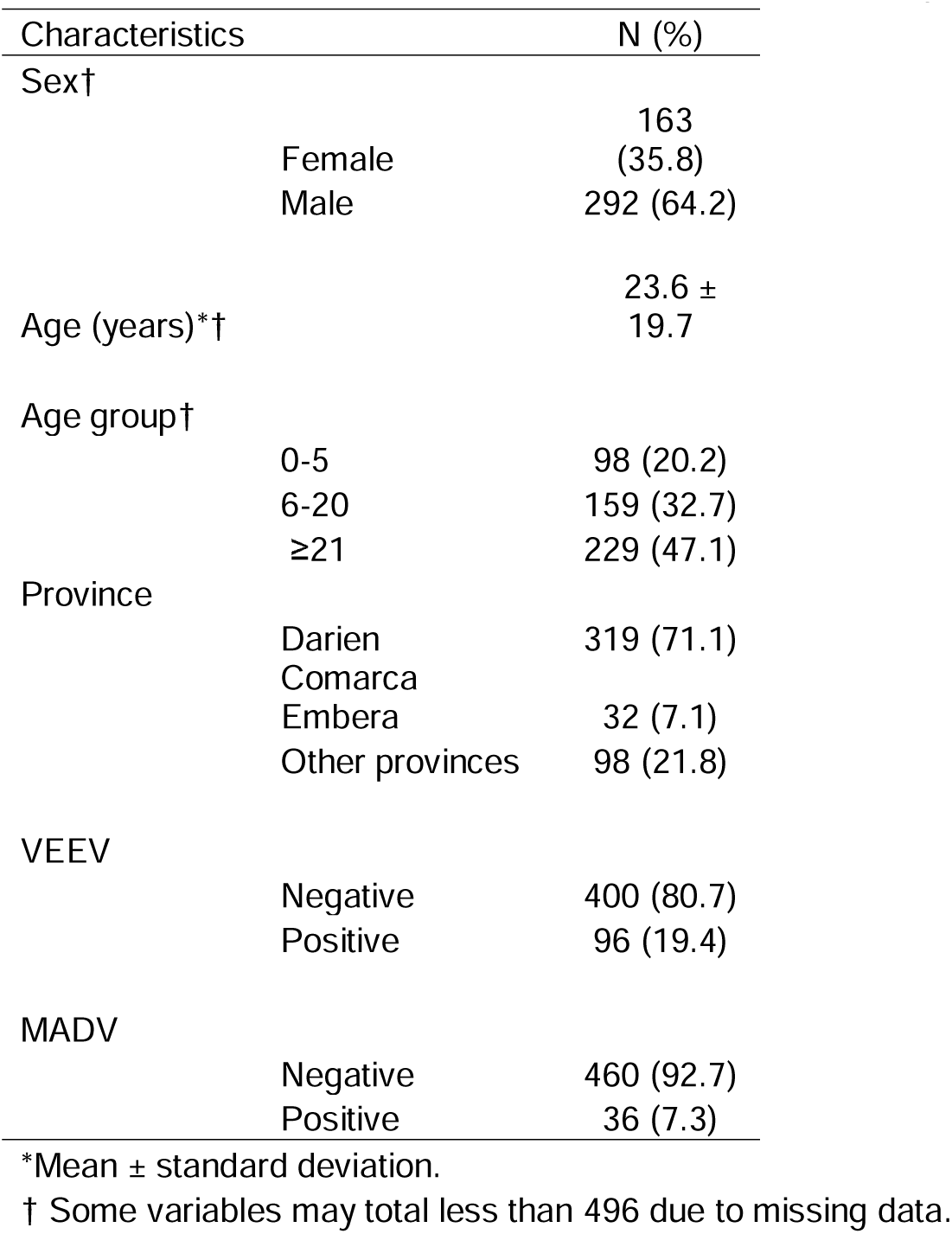
Socio-demographic characteristics of cases for whom samples were submitted to GMI for encephalitic alphavirus testing from 1961 to 2023 (n=496) †.

All human MADV infections were reported from the Darien province. VEEV infections were reported throughout Panama, but most reports (63.4%) were also from the Darien Province (Figure 3). The peak of MADV cases occurred during the 2010 outbreak in the Darien Province, registering 13 laboratory-confirmed cases. The highest number of VEEV cases occurred in 2015 (n=28) (Figure 2). Among the MADV cases, 23 were male and 13 were female. Three cases exhibited mild disease, 11 were moderate, and 17 had severe manifestations, resulting in a mild-to-severe ratio of 3:17. Only 56 VEEV infections had recorded sex information, with a distribution of 39 male and 17 female cases. Severity assessment was only possible in 45 VEEV cases due to incomplete clinical data, of which 10 cases (22.2%) were classified as mild, 25 (55.6%) as moderate, and 10 (22.2%) as severe, resulting in a mild to severe ratio of 1:1. One MADV fatality (1/36, 2.8%), and 8 VEEV fatalities (8/95, 8.4%) were reported.

**Figure 3.**
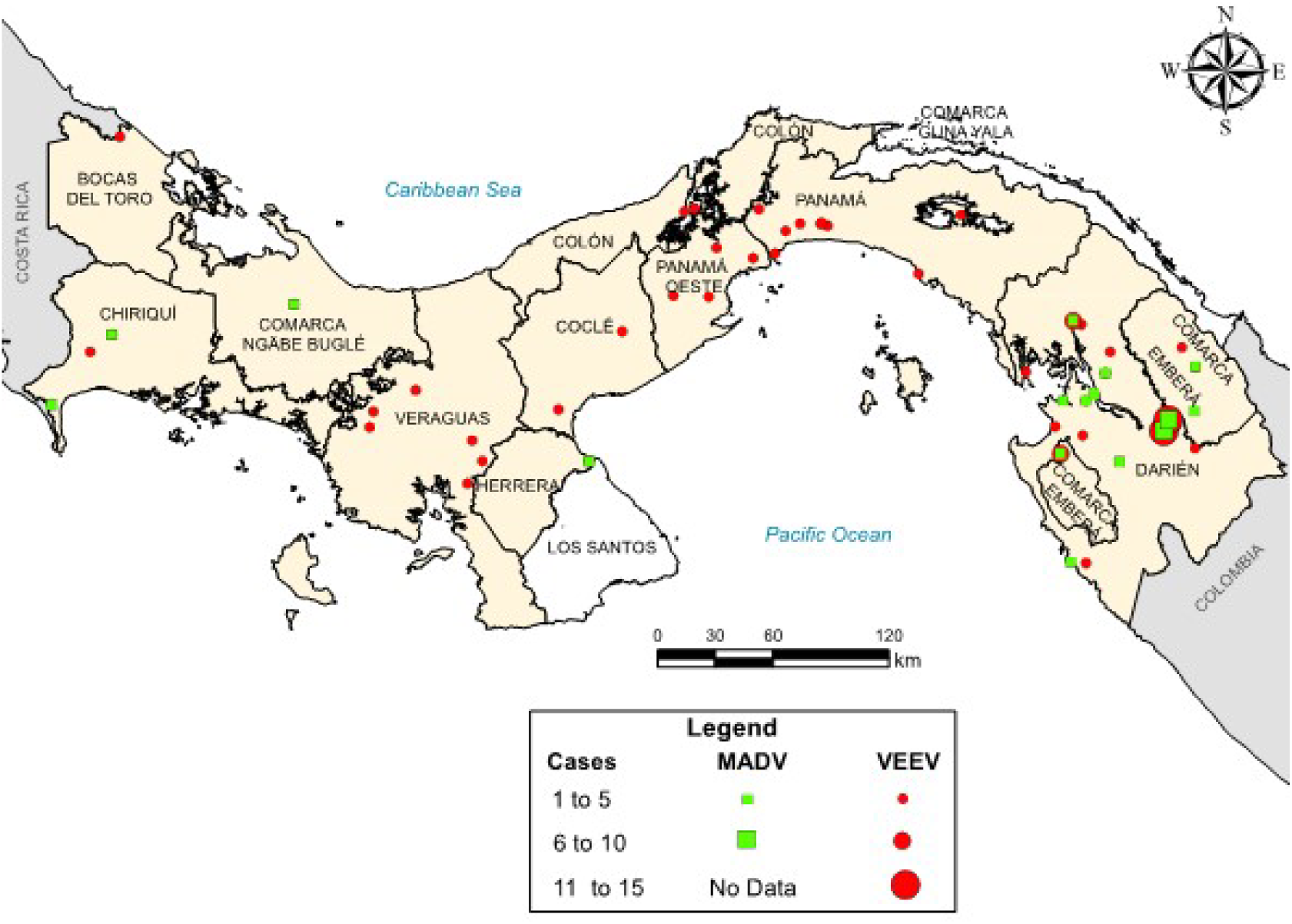
Map of recorded MADV and VEEV cases in Panama. Map showing the distribution of all MADV (green squares) and VEEV (red circles) cases reported in Panama from 1961-2023. MADV cases were only reported in the eastern Panama region, in the province of Darien. MADV cases detected outside Darien, in Chiriquí, Comarca Näbe Bugle and Herrera were reported in members of the border police working in the Darien Province, but at time of symptom onset these cases were detected in their home region.

### MADV and VEEV laboratory testing

We conducted a retrospective analysis to identify the diagnostic methods employed for detecting VEEV and MADV infections from 1961 to 2023. MADV infections were identified nearly exclusively (n=26, 91.9%) by ELISA IgM, with a single case (8.1%) detected by RT-PCR on brain tissue following autopsy. VEEV infections were mostly identified through viral isolation (n=67, 51.1%), followed by ELISA IgM (n=45, 34.3%), and RT-PCR (n=19, 14.5%) (Supplemental Table 2).

### VEEV and MADV clinical presentation

The most frequently documented signs/symptoms of MADV infections included fever (n=29, 81. 0%), neurological symptoms (n=20, 55.6%), headache (n=15, 41.7%), and vomiting (n=15, 41.7%). Fever (n=76, 87.3%), headache (n=51, 58.6%), and vomiting (n=27, 28.1%), were predominantly present in VEEV positive cases. Overall, neurological symptoms were more common in MADV infections, and slightly more common among males. Less common signs/symptoms, including diarrhea (n=8, 9.2%), pharyngitis (n=2, 2.3%), hemorrhage (n=2, 2.3%), and rash (n=1, 1.1%) were more prevalent in VEEV infections (Figure 4).

**Figure 4.**
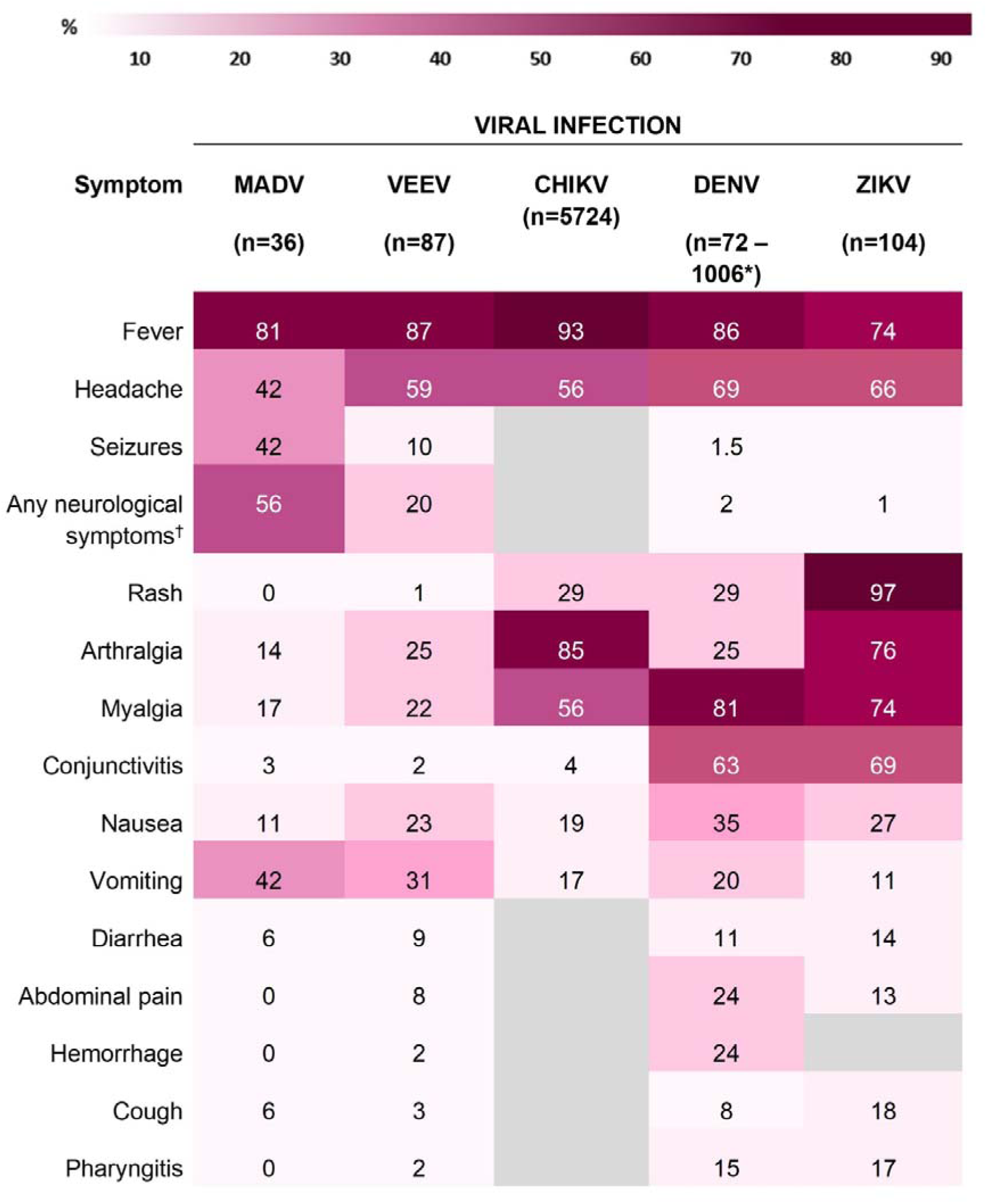
Sign/symptom frequency heatmap by viral infection. Gray blocks denote missing data. * Several datasets from alphavirus cases in Panama and DENV, CHIKV and ZIKV infection cases from Brazil were used to provide more complete symptom data. ^†^ e.g., seizures, focal sensory and/or motor deficits, and diminished level of consciousness.

Fever was consistently reported among both viruses, both sexes and amongst all age groups (Figure 5 and 6). Headaches were also consistently reported in patients infected by both viruses but increased in frequency concurrently with age. In MADV cases, there was an overall higher frequency of neurological symptoms consistently seen in the 0-5 and 6-20 age groups, which contrasts with VEEV infections, where neurological symptoms were reported in the >5-year-old age group. The frequency of neurological symptoms was also higher in males for MADV infections (61% for males vs 45% for females), but equally distributed among VEEV cases (20% for males vs. 19% for females) (Figure 5). Myalgias, arthralgias, and nausea were more commonly seen in VEEV cases, and their frequency also increased with age, with the highest frequency reported in the ≥21-year-old age group (Figure 6). Abdominal pain was only reported amongst VEEV cases and was more commonly reported in females and exclusively reported in the ≥21-year-old age group. Conjunctivitis was exclusively seen in the ≥21-year-old age group for MADV infections. Interestingly, diarrhea was equally distributed among VEEV cases of both sexes until age 20, with only male cases reporting diarrhea in the ≥21-year-old age group.

**Figure 5.**
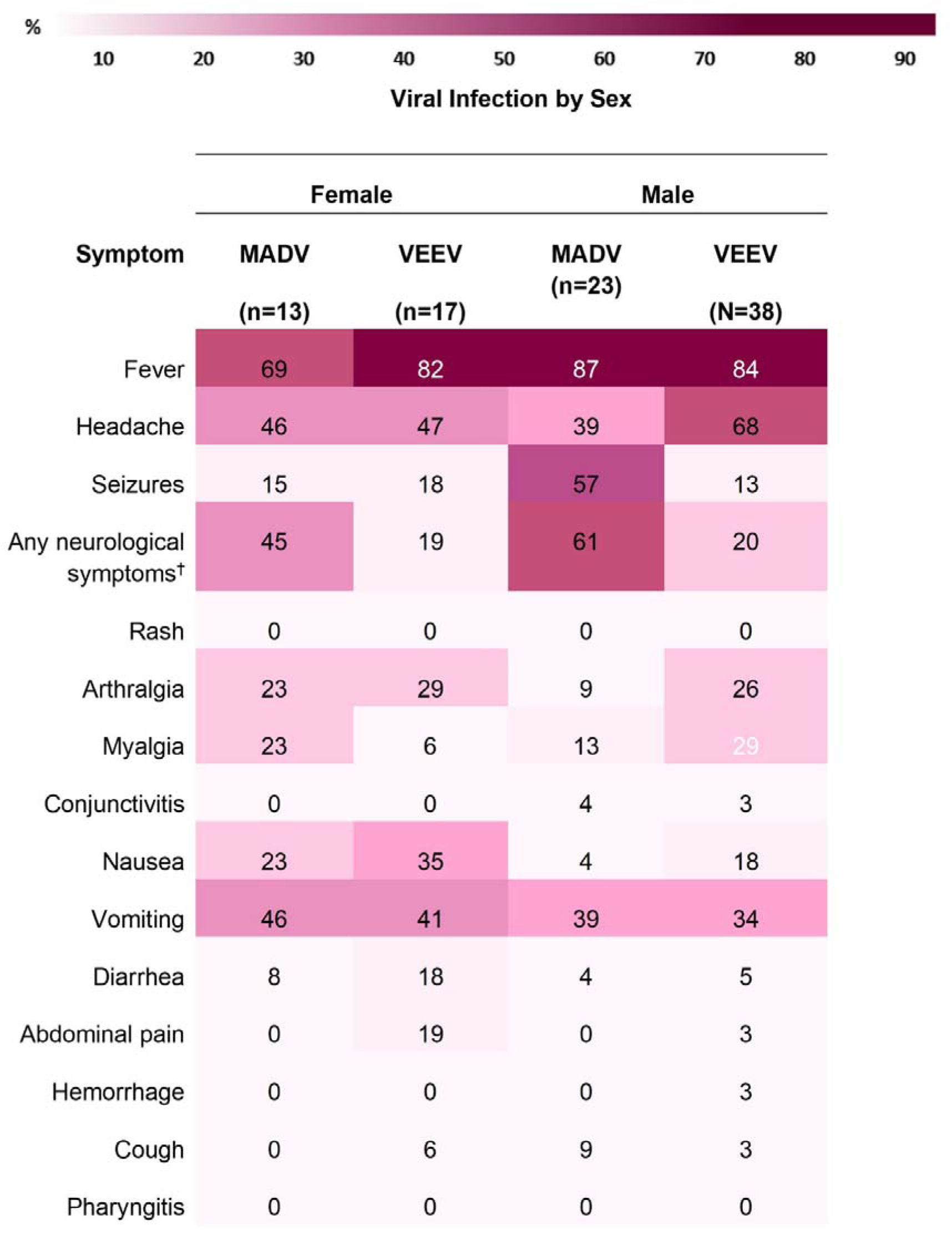
Sign/symptom frequency heatmap by sex and viral infection. ^†^ e.g., seizures, focal sensory and/or motor deficits, and diminished level of consciousness.

**Figure 6.**
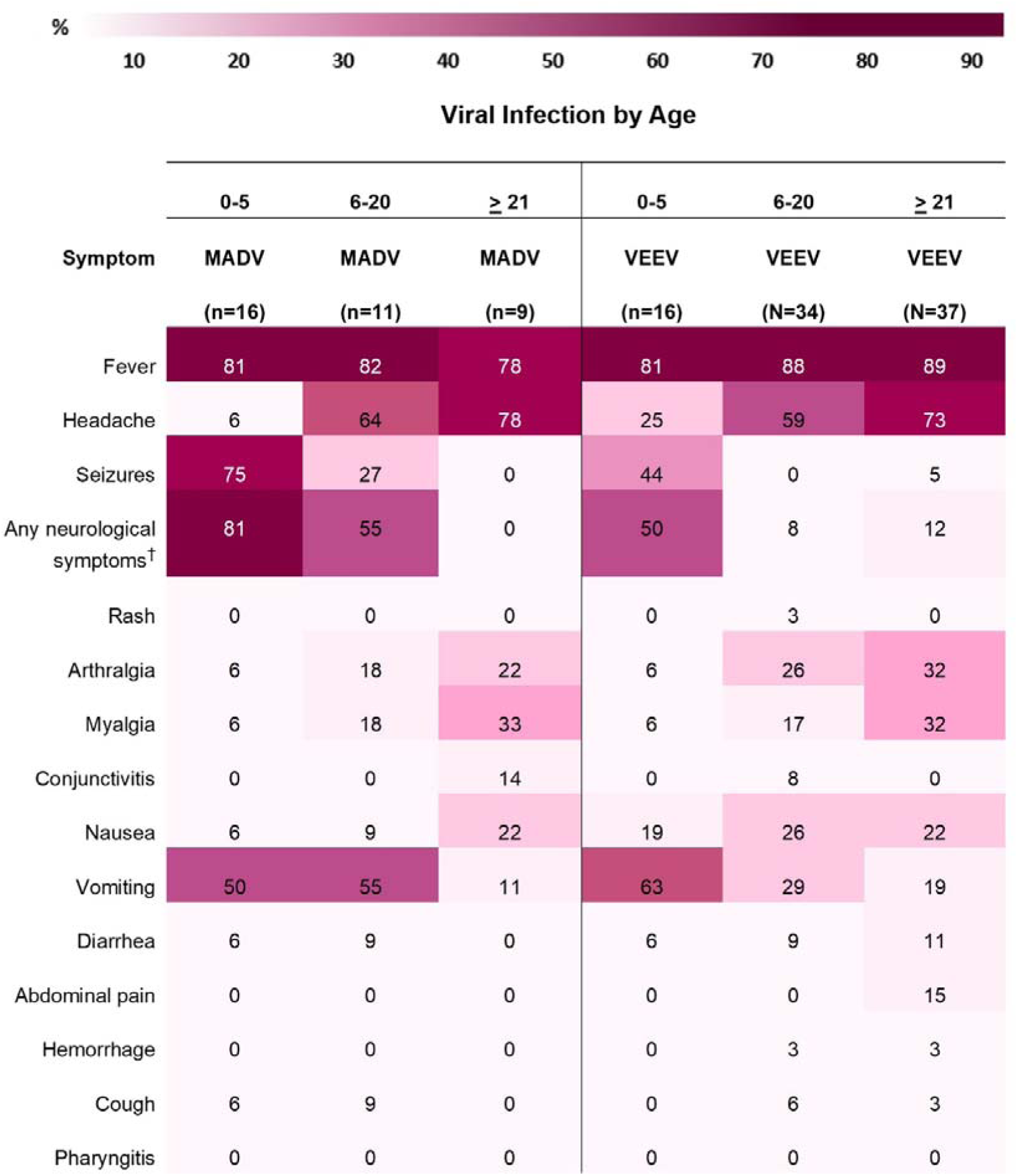
Sign/symptom frequency heatmap by age and viral infection. ^†^ e.g., seizures, focal sensory and/or motor deficits, and diminished level of consciousness.

Logistic regression analysis controlling for sex and age showed that seizures and vomiting were associated with MADV infections to a greater extent than VEEV infections (Supplemental Table 3). At the multivariable level, after variable selection processes, only seizures remained statistically significant when comparing MADV and VEEV (Figure 7A, Supplemental Table 3).

**Figure 7.**
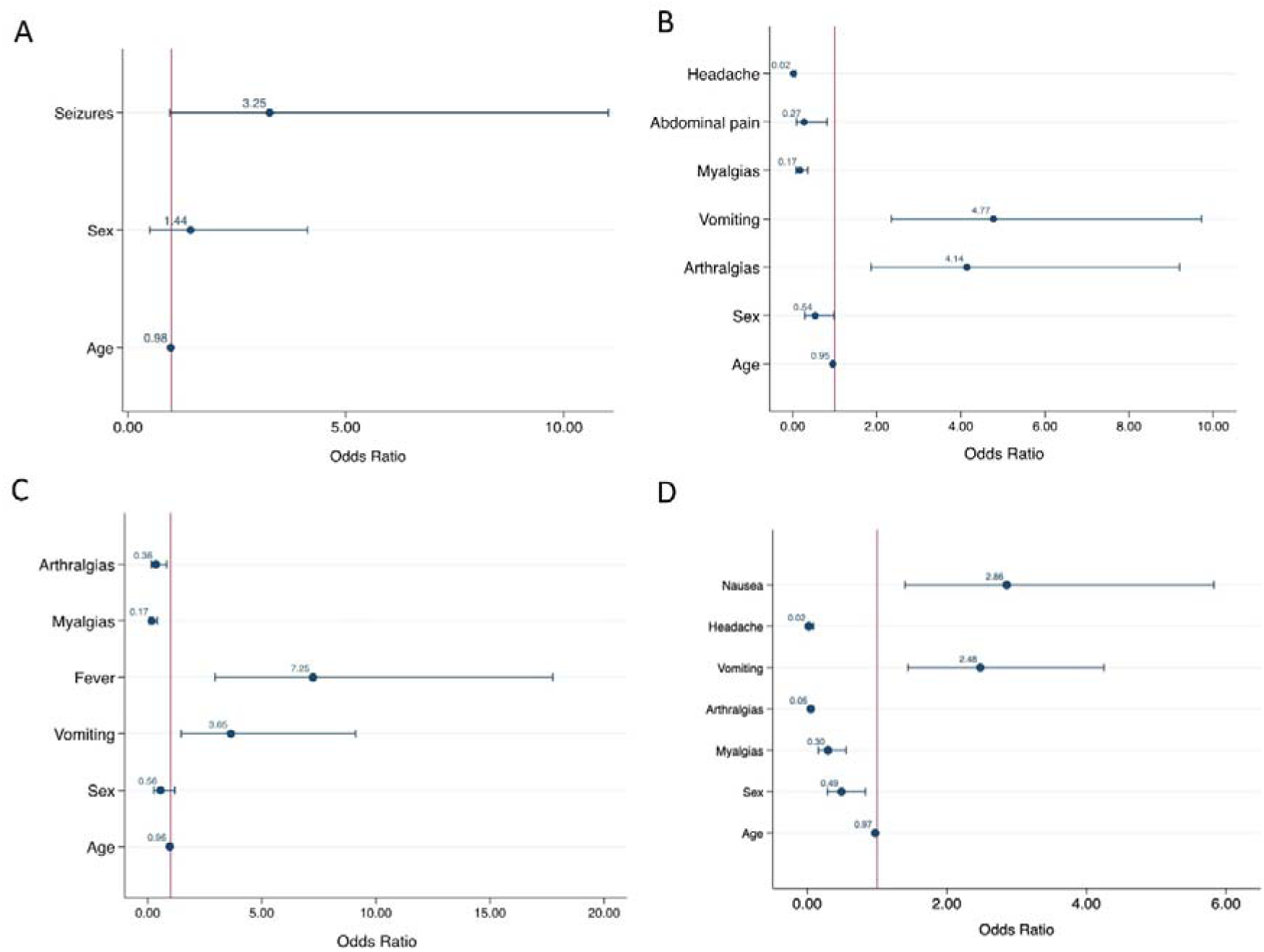
Multivariable logistic regression analysis of associated symptoms of A) MADV vs VEEV infection, and combined encephalitic alphavirus infection versus, B) DENV infection, C) ZIKV infection and D) CHIKV infection. The red vertical line represented number 1, an odds ratio of 1 indicates that the odds of the event are the same in both groups.

### Encephalitic alphavirus versus DENV infection

When alphavirus infection was compared to DENV infection at the univariate level, patients with MADV and VEEV infections were less likely to present with abdominal pain, mucosal bleeding, headaches, myalgias or nausea, while respiratory symptoms, seizures, vomiting, and diarrhea were more frequent in alphavirus infections (Supplemental Table 4).

At the multivariable level, abdominal pain, headache, and myalgias remained less frequent in alphavirus compared to dengue infection, while arthralgia and vomiting remained more frequently associated with alphavirus infection (Figure 7B).

### Encephalitic alphavirus versus ZIKV infection

When alphavirus infection was compared with ZIKV infection at the univariate level, patients with MADV and VEEV infections were less likely to present with respiratory symptoms, pharyngitis, headaches, arthralgias, myalgias, and conjunctivitis, while fever, seizures, and vomiting were more frequent (Supplemental Table 5). At the multivariable level, arthralgia and myalgias were less frequent in alphavirus infection, while fever and vomiting were more common (Figure 7C).

### Encephalitic alphavirus versus CHIKV infection

Univariate analysis of alphavirus infection compared with CHIKV infection showed that fever, respiratory symptoms, headaches, arthralgias, myalgias were less common in encephalitic alphavirus infection, while pharyngitis, diarrhea, and vomiting were more common (Supplemental Table 6). At the multivariable level, arthralgia and myalgia were less frequent in alphavirus infection, and nausea and vomiting were more common (Figure 7D).

## Discussion

In this epidemiological study, we have provided a comprehensive assessment of VEEV and MADV cases in Panama. We summarize and contextualize the clinical findings of human cases of MADV and VEEV in Panama, and identified symptoms that could be used as suggestive of MADV and VEEV infection when compared to other endemic arboviral infections in the region. We have shown that MADV and VEEV cases disproportionally affected males, and MADV occurs more often in children while most VEEV cases occur in adults.

It is unclear if the sex or age-related susceptibility differences of VEEV and MADV are due to a lack of pre-existing immunity, or different exposure risks (e.g., occupational). VEEV has been present in Panama since the mid-20th century when the first human isolate was made in 1961 (16). The first human outbreak of VEEV in Panama occurred in 1967 in U.S soldiers training on the western shores of Gatun Lake (17). Since then, outbreaks of VEEV have been reported periodically in humans. While equine cases of MADV have been documented in Panama since 1936 (18), instances of human cases were infrequent before 2010, despite active human surveillance during outbreaks and widespread mosquito isolations (10, 19, 20).

A 2012 study on the MADV and VEEV seropositivity in humans demonstrated an increasing prevalence of antibodies for VEEV with age demonstrating that the virus is endemic in the region (21). This same trend was not observed for MADV suggesting that the virus recently emerged in humans during the 2010 outbreak. MADV may have gained human virulence since 2010 (8) which may explain why we continue to see human cases. Children may be more susceptible to MADV due to a lack of pre-existing immunity or an immature immune system. The primary risk factors for human exposure to both viruses were found to be farming and fishing (21). Perhaps males spend more time outside performing these activities which puts them at an increased risk for exposure to infected mosquitoes. Our results highlight the need for continued surveillance for VEEV and MADV to better understand these differences.

The Darien province in Panama is a hotspot for VEEV and MADV activity, especially for more recent outbreaks. All MADV human infections have occurred in this region whereas VEEV infections have occurred throughout Panama. Darien is a remote region in eastern Panama near the Colombian border that is inhabited primarily by Indigenous communities. This region contains swamps and forest habitats that can support the enzootic transmission cycle of VEEV and MADV involving rodents and mosquitoes. The Darien region also has a high number of refugee and migrant crossings with over 500,000 reported by the United Nations in 2023 (22). Human migration through this region could result in more cases and the potential spread to other regions. This is highlighted by the MADV cases detected outside Darien, which were reported by members of the border police working in the Darien Province, but symptoms did not develop until they returned to their home regions. While our study reports 168 confirmed human cases of encephalitic alphavirus infection in Panama, the true burden of disease is likely underestimated which is highlighted by the recent finding that 11.9% of dengue-like disease patients were VEEV infections (23).

As described previously throughout the range of enzootic strains, the majority of human VEEV infections in Panama were symptomatic, and reported symptoms included fever, headache, chills, and arthralgia (10). We observed similar symptoms in our study where fever, headache, nausea, and vomiting were frequently reported. Seizures occurred in 10% of cases which is similar to previous reports [reviewed in (24, 25)]. During the 2010 outbreak of MADV, reported symptoms included fever, headache, vomiting, and diarrhea followed by progression to a neurological disease similar to that observed for EEEV (8). The current study found that fever, vomiting, and seizure were the most reported symptoms. Seizures occurred in 42.8% of cases which is significantly higher compared to 10% for VEEV. Most encephalitic alphavirus infections present as a self-limiting febrile illness, but neurological sequelae following recovery from alphavirus infection has been documented at high rates for VEEV and EEEV (50-90%), with individuals reporting one or more long-term symptoms (5, 25). Interestingly, these individuals did not recall having encephalitis or severe neurological signs/symptoms. This suggests that long-term neurological sequelae occur even after mild to moderate clinical presentations. Therefore, the development of any signs of neurologic disease or sequelae should be an important indication of encephalitic alphavirus infection and monitored closely.

In our study vomiting was positively associated with encephalitic alphavirus infection when compared to other arboviruses. Vomiting can occur due to elevated intracranial pressure, which in turn is seen in acute viral encephalitis (26). Thus, vomiting in alphavirus infection may raise concerns for neuroinvasive disease. Additional symptoms positively associated with alphavirus infection was arthralgia when compared to DENV, fever when compared to ZIKV, and nausea when compared to CHIKV. These subtle differences highlight the challenges for distinguishing between the arboviral infections based on clinical presentation alone. Collectively, vomiting and the development of neurological symptoms and seizures seem to be the most predictive symptoms of an encephalitic alphavirus infection when compared to those occurring from other arbovirus infections endemic to Panama. However, a major concern is that by the time a patient presents with neurological symptoms or seizures they are already at risk for the development of severe encephalitic disease. This highlights the need for improved diagnostic tools for the early detection of alphavirus infections.

Our study has several limitations. First, clinical information on alphavirus infections were documented using forms that might not capture detailed clinical and laboratory parameters for both VEEV and MADV infections, as well as incomplete clinical data. Second, it is crucial to note that encephalitic alphavirus cases often occur in rural or remote areas with limited healthcare systems and resources, which could underestimate the number of cases. Third, the limited sample size could impact the statistical power and conclusions, particularly for less frequent symptoms. Fourth, the clinical outcome could be virus strain-dependent and thus vary geographically. Finally, symptoms of encephalitic alphavirus infection were compared with those of DENV, ZIKV, and CHIKV infections from Brazilian cohorts. Although the genetic background and social conditions in Panama may differ from those in Brazil, symptoms of DENV, ZIKV, and CHIKV infections appear to be similar across different populations (27-30).

In summary, outbreaks of MADV and VEEV are expected to continue highlighting the importance for continued surveillance efforts in Panama and other parts of Central and South America. Our findings could serve as a valuable tool for clinical and epidemiological decision-making in regions characterized by endemic arboviral circulation and limited laboratory capacity. Human cases of febrile illness reporting vomiting, nausea, arthralgia, and fever may help indicate acute alphavirus infection and should be monitored closely for signs of neurological disease requiring prompt medical attention.

## Biographical Sketch

Dr. Luis Felipe Rivera, a primary care physician at the Gorgas Memorial Institute in Panama City, specializes in arbovirus surveillance and outbreak response. His primary focus includes diagnosing febrile diseases of unknown origin, tropical medicine, and conducting real time outbreak investigations.

Mr. Carlos Lezcano-Coba, an epidemiologist and virologist at the Gorgas Memorial Institute, is dedicated to arbovirus research. His primary interests lie in the epidemiology and ecology of emerging zoonotic diseases.

## Data Availability

All data produced in the present work are contained in the manuscript.

## Acknowledgments

We wish to express appreciation to Betsy Dutary, Julio Cisneros, Evelia Quiroz and Mariana Garcia who worked to develop laboratory dengue surveillance system and alphavirus surveillance at GMI. We also thanks Kathryn Hanley from New Mexico State University for support with study design and funding. AV, LA and SL are members of the Sistema Nacional de Investigation (SNI), SENACYT, Panama. The views expressed in this work reflect the results of research conducted by the authors and do not necessarily reflect the official policy or position of the Department of Defense, the Navy, or the United States Government. DRS is a U.S. Government employee. This work was prepared as part of her official duties. Title 17, U.S.C., § 105 provides that copyright protection under this title is not available for any work of the U.S. Government. Title 17 U.S.C., § 101 defines a U.S. Government work as work prepared by a military Service member or employee of the U.S. Government as part of that person’s official duties.

