## Supplemental Material for "Clinical and epidemiological characteristics of Madariaga and Venezuelan equine encephalitis virus infections"

**Supplementary laboratory procedures**

**Alphavirus diagnostic**

**Viral isolation and molecular testing**

Direct viral detection was addressed using serum or cerebrospinal fluid (CSF) obtained from patients during the early acute stage of the disease (duration of symptoms, ≤3 days), was used for viral RNA extraction using QIaAmp RNA viral extraction kit (Qiagen, Valencia, CA), and attempt viral isolation in Vero cells supplemented with 2% fetal bovine serum and 1% penicillin/streptomycin. Cells were observed daily for evidence of cytopathic effects (CPE) and samples were passed twice for CPE confirmation. Brain tissues from fatal cases were using to prepare a 10% brain suspension using MEM and 10% FBS and used for viral RNA extraction and attempt viral isolation as described before. Viral RNA from serum, cell supernatant and brain suspension were tested using an alphavirus universal reverse transcription-polymerase chain reaction (RT-PCR) assay or specific MADV/VEEV Real Time RT-PCR as described previously (1).

**Serology**

Acute serum or cerebrospinal fluid (CSF) samples collected during the middle acute phase (duration of symptoms, ≤7) was tested for MADV or VEE virus IgM and IgG antibodies for paired samples, using an enzyme-linked immunosorbent assay as described previously (2, 3). In summary, ELISA antigens were prepared using the TC-83 VEEV and EEEV (prepared by Dr. Robert Shope at the Yale Arbovirus Research Unit in August 1989) strain-infected mouse brain using the sucrose-acetone method. ELISA positive samples by means of IgM or IgG were confirmed using a plaque reduction neutralization test (PRNT), with 80% of reduction as cutoff value. Strains VEEV TC-83 and MADV GML-267113 were used for PRNT confirmation.

**Dengue, Zika and Chikungunya virus infections**

Patients with suspected DENV infection were recruited at the Hospital of the Faculdade de Medicina de São José do Rio Preto, in São Paulo. After initial care was provided, the suspected cases were reported in SINAN (Brazilian notification system). Clinical information of DENV infections was obtained from computerized medical records and disease notification forms according to the WHO 2009 guidelines and the Brazilian Ministry of Health. Blood samples were collected and subjected to relevant diagnostic assays and stored at -80˚C until they were repurposed for additional testing relevant to this study. Dengue was classified as: (i) dengue without warning signs (DwWS), (ii) dengue with warning signs (DWS), or (iii) severe dengue disease (SDD).

**Supplemental Figures and Tables:**

Supplemental Figure 1. Flow chart of alphavirus and endemic arbovirus infections included in this study.

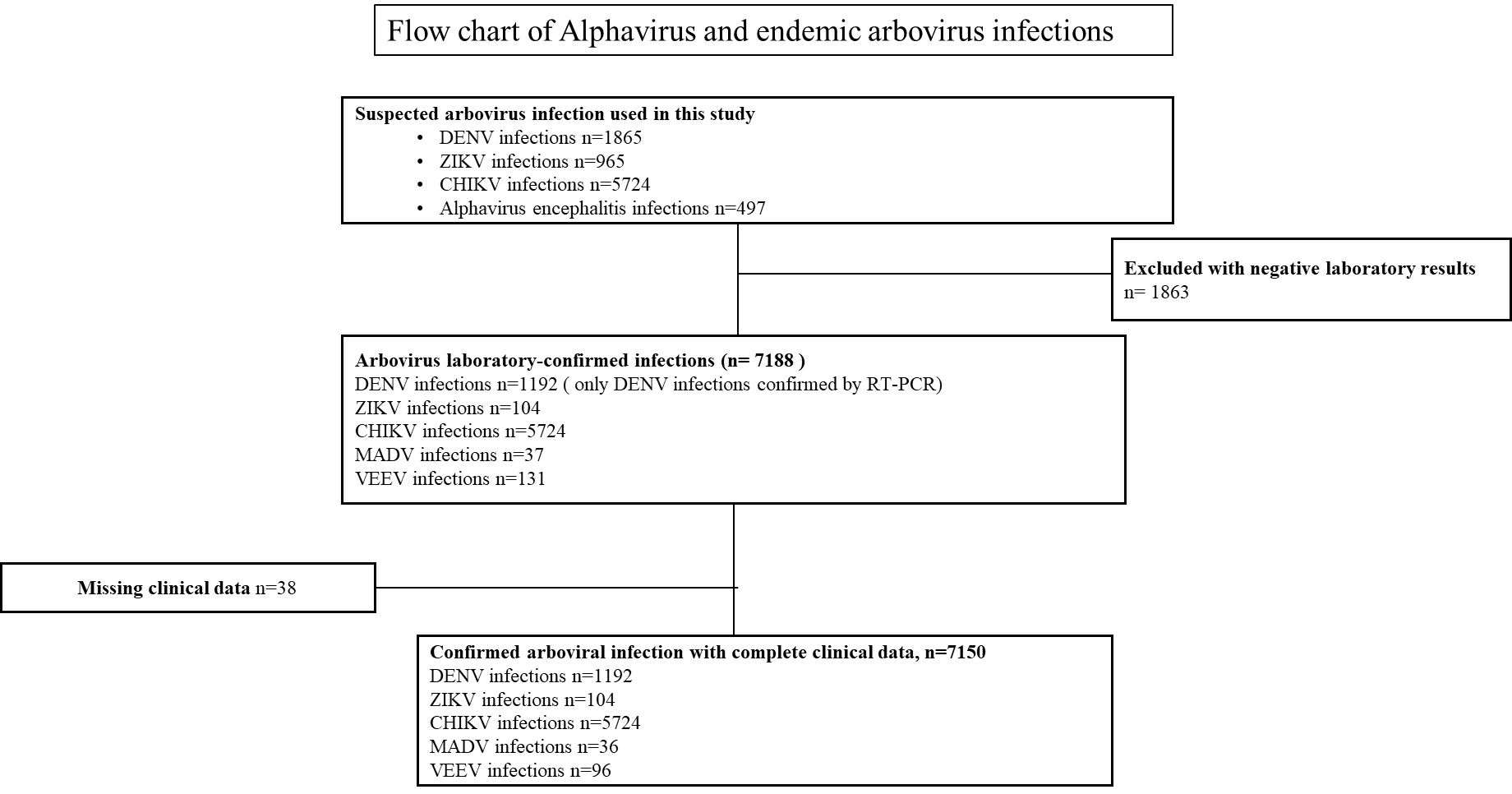

Supplemental Table 1. Kaiser-Meyer-Olkin measure of sampling adequacy for the selected symptom variables, Principal Component Analysis (PCA).

| Variable | Kaiser-Meyer-Olkin |
| --- | --- |
| Abdominal pain | 0.5619 |
| Fever | 0.6824 |
| Respiratory symptoms** | 0.6811 |
| Mucosal bleeding | 0.5275 |
| Seizures | 0.6531 |
| Pharyngitis | 0.6648 |
| Headache | 0.7039 |
| Arthralgias | 0.6224 |
| Nausea | 0.6635 |
| Myalgias | 0.6939 |
| Rash | 0.7134 |
| Conjunctivitis | 0.6632 |
| Vomiting | 0.6008 |
| Diarrhea | 0.7039 |
| Overall | 0.6554 |

Supplemental Table 2. Distribution of MADV and VEEV cases by year, diagnostic method, and detection type.

| **Year** | **MADV positive cases** | **MADV Diagnostic Methods** | **MADV Detection Method** | **VEEV positive cases** | **VEEV Diagnostic Methods** | **VEEV Detection Method** |
| --- | --- | --- | --- | --- | --- | --- |
| 1961 | 0 |  |  | 1 | Viral isolation | Arbovirus surveillance |
| 1962 | 0 |  |  | 3 | Viral isolation | Arbovirus surveillance |
| 1964 | 0 |  |  | 1 | Viral isolation | Arbovirus surveillance |
| 1967 | 0 |  |  | 7 | Viral isolation | Arbovirus surveillance |
| 1973 | 0 |  |  | 10 | Viral isolation | Arbovirus surveillance |
| 1977 | 0 |  |  | 9 | Viral isolation | Arbovirus surveillance |
| 1981 | 0 |  |  | 1 | Viral isolation | Arbovirus surveillance |
|  |  |  |  | 5 | ELISA IgM | Arbovirus surveillance |
| 1990 | 0 |  |  | 1 | Viral isolation | Arbovirus surveillance |
| 1991 | 0 |  |  | 1 | Viral isolation | Arbovirus surveillance |
| 1993 | 0 |  |  | 1 | Viral isolation | Arbovirus surveillance |
| 1995 | 0 |  |  | 2 | Viral isolation | Arbovirus surveillance |
| 1996 | 0 |  |  | 1 | Viral isolation | Arbovirus surveillance |
| 1997 | 0 |  |  | 2 | Viral isolation | Arbovirus surveillance |
| 1998 | 0 |  |  | 7 | Viral isolation | Arbovirus surveillance |
| 2000 | 0 |  |  | 1 | Viral isolation | Arbovirus surveillance |
| 2001 | 0 |  |  | 6 | Viral isolation | Arbovirus surveillance |
| 2002 | 0 |  |  | 1 | Viral isolation | Arbovirus surveillance |
| 2003 | 0 |  |  | 5 | Viral isolation | Arbovirus surveillance |
| 2004 | 0 |  |  | 3 | Viral isolation | Arbovirus surveillance |
| 2010 | 14 | ELISA IgM | Outbreak | 8 | ELISA IgM | Outbreak |
|  |  |  |  | 1 | RT-PCR | Outbreak |
|  |  |  |  | 3 | Viral isolation | Outbreak |
| 2015 | 5 | ELISA IgM | Outbreak | 13 | RT-PCR | Outbreak |
|  |  |  |  | 15 | ELISA IgM | Outbreak |
| 2016 | 3 | ELISA IgM | Outbreak |  |  |  |
| 2017 | 1 | RT-PCR | Arbovirus surveillance | 10 | ELISA IgM | Outbreak |
|  | 6 | ELISA IgM | Outbreak |  |  |  |
| 2019 | 6 | ELISA IgM | Outbreak | 7 | ELISA IgM | Outbreak |
|  |  |  |  | 1 | Viral isolation | Outbreak |
| 2021 |  |  |  | 3 | RT-PCR | Arbovirus surveillance |
| 2023 | 2 | RT-PCR | Arbovirus surveillance | 2 | RT-PCR | Outbreak |

Supplemental Table 3. Symptoms associated with MADV cases compared to VEEV controls (n=496) †.

| Characteristics | | Univariate analysis | | | Multiple regression, nested model | | |
| --- | --- | --- | --- | --- | --- | --- | --- |
|  |  | OR | CI 95% | p | OR | 95% CI | p |
| Abdominal pain | |  |  |  |  |  |  |
|  | No | Ref. |  |  | Ref. |  |  |
|  | Yes | 0.78 | 0.08 - 7.32 | 0.832 |  |  |  |
| Fever | |  |  |  |  |  |  |
|  | No | Ref. |  |  | Ref. |  |  |
|  | Yes | 0.72 | 0.23 - 2.29 | 0.582 |  |  |  |
| Respiratory symptoms** | |  |  |  |  |  |  |
|  | No | Ref. |  |  | Ref. |  |  |
|  | Yes | 3.12 | 0.59 - 16.39 | 0.180 |  |  |  |
| Mucosal bleeding** | |  |  |  |  |  |  |
|  | No | Ref. |  |  | Ref. |  |  |
|  | Yes | Empty |  |  |  |  |  |
| Seizures | |  |  |  |  |  |  |
|  | No | Ref. |  |  | Ref. |  |  |
|  | Yes | 7.49 | 2.75 - 20.46 | **<0.001** | 3.25 | 0.96 - 11.02 | 0.058 |
| Pharyngitis** | |  |  |  |  |  |  |
|  | No | Ref. |  |  | Ref. |  |  |
|  | Yes | Empty |  |  |  |  |  |
| Headache | |  |  |  |  |  |  |
|  | No | Ref. |  |  | Ref. |  |  |
|  | Yes | 0.31 | 0.07 - 1.40 | 0.127 |  |  |  |
| Arthralgias | |  |  |  |  |  |  |
|  | No | Ref. |  |  | Ref. |  |  |
|  | Yes | 0.65 | 0.19 - 1.98 | 0.415 |  |  |  |
| Nausea | |  |  |  |  |  |  |
|  | No | Ref. |  |  | Ref. |  |  |
|  | Yes | 0.66 | 0.20 - 2.13 | 0.485 |  |  |  |
| Myalgias | |  |  |  |  |  |  |
|  | No | Ref. |  |  | Ref. |  |  |
|  | Yes | 0.92 | 0.31 - 2.76 | 0.88 |  |  |  |
| Rash** | |  |  |  |  |  |  |
|  | No | Ref. |  |  | Ref. |  |  |
|  | Yes | Empty |  |  |  |  |  |
| Conjunctivitis** | |  |  |  |  |  |  |
|  | No | Ref. |  |  | Ref. |  |  |
|  | Yes | 3.25 | 0.19 - 53.66 | 0.410 |  |  |  |
| Vomiting | |  |  |  |  |  |  |
|  | No | Ref. |  |  | Ref. |  |  |
|  | Yes | 2.37 | 1.01 - 5.59 | **0.049** |  |  |  |
| Diarrhea | |  |  |  |  |  |  |
|  | No | Ref. |  |  | Ref. |  |  |
|  | Yes | 0.78 | 0.16 - 3.89 | 0.760 |  |  |  |
| *Adjusted by age and sex. | | | | | | | |
| **Variables excluded from the multivariable model due to missing values. | | | | | |  |  |
| † Some variables may total less than 9644 due to missing data. | | | | |  |  |  |
| **OR:** Odds Ratio. 95% CI: 95% Confidence Interval. |  |  |  |  |  |  |  |

Supplemental Table 4. Symptoms associated with alphavirus cases with DENV controls (n=9644) †.

| Characteristics | | Univariate analysis | | | Multiple regression, nested model | | |
| --- | --- | --- | --- | --- | --- | --- | --- |
|  |  | OR | 95% CI | p | OR | 95% CI | p |
| Abdominal pain | |  |  |  |  |  |  |
|  | No | Ref. |  |  | Ref. |  |  |
|  | Yes | 0.19 | 0.07 - 0.48 | **<0.001** | 0.27 | 0.09 - 0.82 | **0.021** |
| Fever | |  |  |  |  |  |  |
|  | No | Ref. |  |  | Ref. |  |  |
|  | Yes | 1.08 | 0.63 - 1.85 | 0.768 |  |  |  |
| Respiratory symptoms | |  |  |  |  |  |  |
|  | No | Ref. |  |  | Ref. |  |  |
|  | Yes | 5.08 | 1.89 - 13.65 | **0.001** |  |  |  |
| Mucosal bleeding | |  |  |  |  |  |  |
|  | No | Ref. |  |  | Ref. |  |  |
|  | Yes | 0.20 | 0.04 - 0.80 | **0.024** |  |  |  |
| Seizures** | |  |  |  |  |  |  |
|  | No | Ref. |  |  | Ref. |  |  |
|  | Yes | 43.92 | 17.40 - 110.85 | **<0.001** |  |  |  |
| Pharyngitis** | |  |  |  |  |  |  |
|  | No | Ref. |  |  | Ref. |  |  |
|  | Yes | 1.65 | 0.36 - 7.47 | 0.514 |  |  |  |
| Headache | |  |  |  |  |  |  |
|  | No | Ref. |  |  | Ref. |  |  |
|  | Yes | 0.12 | 0.07 - 0.21 | **<0.001** | 0.01 | 0.01 - .07 | **<0.001** |
| Arthralgias | |  |  |  |  |  |  |
|  | No | Ref. |  |  | Ref. |  |  |
|  | Yes | 0.81 | 0.50 - 1.31 | 0.399 | 4.14 | 1.86 - 9.20 | **<0.001** |
| Nausea | |  |  |  |  |  |  |
|  | No | Ref. |  |  | Ref. |  |  |
|  | Yes | 0.5 | 0.30 - 0.81 | **0.005** |  |  |  |
| Myalgias | |  |  |  |  |  |  |
|  | No | Ref. |  |  | Ref. |  |  |
|  | Yes | 0.09 | 0.05 - 0.15 | **<0.001** | 0.17 | 0.08 - 0.36 | **<0.001** |
| Rash** | |  |  |  |  |  |  |
|  | No | Ref. |  |  | Ref. |  |  |
|  | Yes | empty |  |  |  |  |  |
| Conjunctivitis** | |  |  |  |  |  |  |
|  | No | Ref. |  |  | Ref. |  |  |
|  | Yes | 0.42 | 0.10 - 1.78 | 0.245 |  |  |  |
| Vomiting | |  |  |  |  |  |  |
|  | No | Ref. |  |  | Ref. |  |  |
|  | Yes | 2.17 | 1.44 - 3.27 | **<0.001** | 4.77 | 2.34 - 9.72 | **<0.001** |
| Diarrhea** | |  |  |  |  |  |  |
|  | No | Ref. |  |  | Ref. |  |  |
|  | Yes | 10.64 | 4.33 - 26.13 | **<0.001** |  |  |  |
| *Adjusted by age and sex. | | | | | | | |
| **Variables excluded from the multivariable model due to missing values. | | | | |  |  |  |
| † Some variables may total less than 9644 due to missing data. | | | | |  |  |  |
| **OR:** Odds Ratio. 95% CI: 95% Confidence Interval. |  |  |  |  |  |  |  |

Supplemental Table 5. Symptoms associated with alphavirus cases with ZIKV controls (n=9644) †.

| Characteristics | | Univariate analysis | | | Multiple regression, nested model | | |
| --- | --- | --- | --- | --- | --- | --- | --- |
|  |  | OR | 95%CI | p | OR | 95% CI | p |
| Abdominal pain | |  |  |  |  |  |  |
|  | No | Ref. |  |  | Ref. |  |  |
|  | Yes | 0.43 | 0.15 - 1.25 | 0.124 |  |  |  |
| Fever | |  |  |  |  |  |  |
|  | No | Ref. |  |  | Ref. |  |  |
|  | Yes | 3.70 | 2.00 - 6.83 | **<0.001** | 7.25 | 2.96 - 17.76 | **<0.001** |
| Respiratory symptoms | |  |  |  |  |  |  |
|  | No | Ref. |  |  | Ref. |  |  |
|  | Yes | 0.32 | 0.12 - 0.84 | **0.021** |  |  |  |
| Mucosal bleeding | |  |  |  |  |  |  |
|  | No | Ref. |  |  | Ref. |  |  |
|  | Yes | 2.58 | 0.23 - 28.78 | 0.442 |  |  |  |
| Seizures** | |  |  |  |  |  |  |
|  | No | Ref. |  |  | Ref. |  |  |
|  | Yes | 31.78 | 4.21 - 239.63 | **0.001** |  |  |  |
| Pharyngitis** | |  |  |  |  |  |  |
|  | No | Ref. |  |  | Ref. |  |  |
|  | Yes | 0.12 | 0.02 - 0.51 | **0.005** |  |  |  |
| Headache | |  |  |  |  |  |  |
|  | No | Ref. |  |  | Ref. |  |  |
|  | Yes | 0.17 | 0.09 - 0.31 | **<0.001** |  |  |  |
| Arthralgias | |  |  |  |  |  |  |
|  | No | Ref. |  |  | Ref. |  |  |
|  | Yes | 0.17 | 0.09 - 0.29 | **<0.001** | 0.36 | 0.15 - 0.82 | **0.016** |
| Nausea | |  |  |  |  |  |  |
|  | No | Ref. |  |  | Ref. |  |  |
|  | Yes | 0.76 | 0.41 - 1.42 | 0.395 |  |  |  |
| Myalgias | |  |  |  |  |  |  |
|  | No | Ref. |  |  | Ref. |  |  |
|  | Yes | 0.17 | 0.09 - 0.31 | **<0.001** | 0.17 | 0.07 - 0.42 | **<0.001** |
| Rash** | |  |  |  |  |  |  |
|  | No | Ref. |  |  | Ref. |  |  |
|  | Yes | empty |  |  |  |  |  |
| Conjunctivitis** | |  |  |  |  |  |  |
|  | No | Ref. |  |  | Ref. |  |  |
|  | Yes | 0.02 | 0.01 - 0.07 | **<0.001** |  |  |  |
| Vomiting | |  |  |  |  |  |  |
|  | No | Ref. |  |  | Ref. |  |  |
|  | Yes | 4.79 | 2.41 - 9.51 | **<0.001** | 3.65 | 1.46 - 9.12 | **0.006** |
| Diarrhea | |  |  |  |  |  |  |
|  | No | Ref. |  |  | Ref. |  |  |
|  | Yes | 0.72 | 0.31 - 1.65 | 0.439 |  |  |  |
| *Adjusted by age and sex. | | | | | | | |
| **Variables excluded from the multivariable model due to missing values. | | | | | |  |  |
| † Some variables may total less than 9644 due to missing data. | | | | |  |  |  |
| **OR:** Odds Ratio. 95% CI: 95% Confidence Interval. |  |  |  |  |  |  |  |

Supplemental Table 6. Symptoms associated with alphavirus cases compared to CHIKV controls (n=9644) †.

| Characteristics | | Bivariate analysis | | | Multiple regression, nested model | | |
| --- | --- | --- | --- | --- | --- | --- | --- |
|  |  | OR | 95% CI | p | OR | 95% CI | p |
| Abdominal pain** | |  |  |  |  |  |  |
|  | No | Ref. |  |  | Ref. |  |  |
|  | Yes | Empty |  |  |  |  |  |
| Fever | |  |  |  |  |  |  |
|  | No | Ref. |  |  | Ref. |  |  |
|  | Yes | 0.48 | 0.28 - 0.82 | **0.007** |  |  |  |
| Respiratory symptoms** | |  |  |  |  |  |  |
|  | No | Ref. |  |  | Ref. |  |  |
|  | Yes | 107.68 | 26.58 - 436.25 | **<0.001** |  |  |  |
| Mucosal bleeding** | |  |  |  |  |  |  |
|  | No | Ref. |  |  | Ref. |  |  |
|  | Yes | Empty |  |  |  |  |  |
| Seizures** | |  |  |  |  |  |  |
|  | No | Ref. |  |  | Ref. |  |  |
|  | Yes | Empty |  |  |  |  |  |
| Pharyngitis** | |  |  |  |  |  |  |
|  | No | Ref. |  |  | Ref. |  |  |
|  | Yes | 96.86 | 8.72 - 1075.51 | **<0.001** |  |  |  |
| Headache | |  |  |  |  |  |  |
|  | No | Ref. |  |  | Ref. |  |  |
|  | Yes | 0.14 | 0.08 - 0.22 | **<0.001** | 0.02 | 0.01 - 0.09 | **<0.001** |
| Arthralgias | |  |  |  |  |  |  |
|  | No | Ref. |  |  | Ref. |  |  |
|  | Yes | 0.04 | 0.02 - 0.06 | **<0.001** | 0.05 | 0.03 - 0.09 | **<0.001** |
| Nausea | |  |  |  |  |  |  |
|  | No | Ref. |  |  | Ref. |  |  |
|  | Yes | 0.91 | 0.57 - 1.47 | 0.703 | 2.86 | 1.40 - 5.82 | **0.004** |
| Myalgias | |  |  |  |  |  |  |
|  | No | Ref. |  |  | Ref. |  |  |
|  | Yes | 0.15 | 0.09 - 0.24 | **<0.001** | 0.29 | 0.16 - 0.56 | **<0.001** |
| Rash** | |  |  |  |  |  |  |
|  | No | Ref. |  |  | Ref. |  |  |
|  | Yes | Empty |  |  |  |  |  |
| Conjunctivitis | |  |  |  |  |  |  |
|  | No | Ref. |  |  | Ref. |  |  |
|  | Yes | 0.42 | 0.10 - 1.70 | 0.225 |  |  |  |
| Vomiting | |  |  |  |  |  |  |
|  | No | Ref. |  |  | Ref. |  |  |
|  | Yes | 2.28 | 1.55 - 3.36 | **<0.001** | 2.48 | 1.44 - 4.25 | **0.001** |
| Diarrhea** | |  |  |  |  |  |  |
|  | No | Ref. |  |  | Ref. |  |  |
|  | Yes | 259.55 | 56.22 - 1198.35 | **<0.001** |  |  |  |
| *Adjusted by age and sex. | | | | | | | |
| **Variables excluded from the multivariable model due to missing values. | | | | | |  |  |
| † Some variables may total less than 9644 due to missing data. | | | | |  |  |  |
| **OR:** Odds Ratio. 95% CI: 95% Confidence Interval. |  |  |  |  |  |  |  |
